# State-Level Variability in Implementation Approaches to SNAP Food Restriction Waivers: A Content Analysis of Bills, Waiver Requests, and Waiver Approvals

**DOI:** 10.64898/2026.07.23.26358592

**Authors:** Emily W. Duffy, Miguel Ángel López, Emily Dimond, Laura E. Balis, Elizabeth Piekarz-Porter, Alyssa J. Moran, Carolina Morales Serrano, Elizabeth T. Anderson Steeves

## Abstract

**Background:** Since 2025, US states have rapidly adopted waivers to restrict the purchase of ‘non-nutritious items’ like soda and candy with Supplemental Nutrition Assistance Program (SNAP) benefits. States have proposed various approaches in terms of food categories restricted and implementation strategies, which may influence the downstream effects of waivers on program participation and participants’ diets and health.

**Objective:** To examine and describe contextual information related to the adoption and implementation of state-level SNAP food restriction waivers.

**Design:** A content analysis was conducted including publicly available state-level: (1) bills introduced related to SNAP food restriction waivers (January 2025 to March 2026), (2) waiver requests, and (3) waiver approvals (up to June 2026). Codebooks that captured key elements, such as food and beverage group restrictions and definitions, rationale for restrictions, implementation supports, and evaluation plans, were applied independently by two coders, and discrepancies were resolved through consensus.

**Participants/setting:** United States

**Intervention:** N/A

**Main outcome measures:** N/A

**Statistical analyses performed:** Descriptive statistics were used to summarize the presence of coded elements in documents.

**Results:** Seventy-one bills, 22 waiver requests, and 23 waiver approvals were included. Bills mostly proposed restricting candy (65%) and soft drinks (59%). Of the approved waivers, all restrict the purchase of specific beverage groups with SNAP benefits and 65% restrict the purchase of specific food groups with SNAP benefits. There is a high level of variability in how the same food or beverage groups (e.g., soda) are defined across states. States varied in the implementation supports described in waiver requests, describing strategies like communications plans, nutrition education, and staff training.

**Conclusions:** The current state of SNAP food restriction waivers is highly variable and shifting rapidly. Understanding characteristics of early-adopting states can inform future impact and implementation evaluations of waivers and efforts of additional states that may pursue SNAP food restriction waivers.

## Introduction

The United States Department of Agriculture (USDA) Supplemental Nutrition Assistance Program (SNAP) is the largest federally funded nutrition assistance program in the United States (US), providing support to more than 42 million people in 2025^1^. Historically, SNAP had limited restrictions on the types of foods and beverages that cannot be purchased with program benefits (e.g., prepared foods, alcohol). Waivers to test further food group restrictions (e.g., soda, candy) have been previously requested by states and denied by the USDA. During the past two years, the Make America Healthy Again (MAHA) initiative, led by Health and Human Services Secretary Robert F. Kennedy Jr., has prioritized USDA approval of state waivers for SNAP food and beverage restrictions, marking a major shift in USDA policy^2^. These waivers, also known as pilot or demonstration projects, allow the USDA to temporarily waive SNAP regulations and test changes to SNAP for up to five years. Since May 2025, there has been rapid adoption of SNAP food restriction waivers, with 23 states receiving USDA approval to restrict the use of SNAP for “non-nutritious items like soda and candy”^3^.

Rapid action on SNAP food restriction waivers is continuing on multiple fronts. First, state legislators are introducing bills that urge Governors and/or state agencies to submit waiver requests within a specific timeline. Second, governors and/or state agencies are responding to legislative action by submitting SNAP waiver requests. Third, governors and/or state agencies are unilaterally submitting SNAP waiver requests in the absence of state legislation. It is not a requirement for states to enact waiver-related legislation to receive approval from USDA, but this is one route that states are taking to adopt food restriction waivers. Consequently, state bills provide vital insight into states’ policy goals and interest in pursuing food restriction waivers. Additionally, prior content analyses of bills have noted that factors such as bill sponsors’ party affiliation(s), message framing, and evidence types used to justify government intervention may influence policy adoption^4–6^. Therefore, an initial understanding of how bills are framed may also be informative for policy makers, researchers, or practitioners working on SNAP food restrictions or similar policies such as SNAP healthy food incentives.

States’ waiver requests and approvals provide key insights into how states are planning for waiver implementation. One prior study examined SNAP food restriction waiver requests and approvals in the 12 states with approved waivers as of late 2025 and found considerable variability in states’ implementation approaches, including scope of restricted foods and beverages and evaluation plans^7^. This work, prior evaluations of similar food policies (such as sweetened beverage taxes), and implementation science literature suggest that this waiver variability may influence impact on SNAP participants’ experiences, participation, and dietary intake as well as responses from the food industry. For example, implementation studies of sweetened beverage taxes have demonstrated that factors such as the complexity of the policy, time for implementation preparation, communication and outreach strategies to relevant groups, and resources for staffing influenced the policies’ overall impact on diet-related outcomes^8,9^. Additionally, in June 2026, a federal judge ruled that USDA did not follow its own administrative procedures in the waiver request and approval process and blocked the waivers in Colorado, Iowa, Nebraska, Tennessee, and West Virgina^10^. A comprehensive understanding of plans of early-adopting states, in particular states that adopted prior to the 2026 court ruling, can provide critical information that later adopters can draw on to inform and improve their own waiver adoption and implementation processes^11^, such as definitions of food and beverages, and communication strategies. Moreover, characterizing the context of waiver adoption and state-level variability in waiver implementation can provide valuable information for future evaluations and interventions to support SNAP participants and retailers. Thus, a content analysis of state-level SNAP food restriction waiver-related documents was conducted.

## Methods

### Sample

The sample included three document types: state bills, state waiver requests to USDA, and USDA waiver approvals.

### State Bills

The sample of bills included state bills from all 50 states introduced in the 2025 legislative session and 2026 legislative session through March 2026. Systematic searches were conducted in the Legiscan database using key terms such as “SNAP” and “Supplemental Nutrition Assistance Program”. Bills were included if they referenced requiring the state agency or governor to submit a waiver request to USDA to restrict food groups that could be purchased with SNAP benefits. All search results were documented, and bill summaries and text were reviewed individually by team members to determine inclusion. The final list of included bills was cross-checked against results from other legislative databases: NexisUni, Westlaw, and Quorum.

### USDA Waiver Requests and Approvals

SNAP food restriction waiver requests and approvals were collected from the USDA website for every state with an approved food restriction waiver as of June 2026 (n=22 and 23, respectively). Waiver requests are the documents that state agencies submit to USDA outlining plans for the waiver, such as the food or beverage groups to be restricted, justification for the request, anticipated impacts and implementation, and plans for communication, monitoring, and evaluation. The waiver approvals are documents sent to state agencies from the USDA, outlining the terms and conditions, such as implementation date, length of the pilot, and communication, monitoring, reporting, and evaluation requirements.

### Codebook Development

Codebooks were developed for each document type (i.e., bills, waiver requests, and waiver approvals) (Supplemental File 1). Codebooks were developed based on an initial review of each document type to capture key elements related to waiver adoption and implementation and based on items from similar content analyses^4,12^. Basic information on document types, submission or introduction dates, state, food and beverage groups to be restricted or added, and definitions of food and beverage groups were included in each codebook. Other items were included as appropriate depending on key elements of each document (e.g., bills did not include detailed evaluation plans, so evaluation items were excluded). Related to waiver adoption, key elements such as party affiliation for bill sponsor(s) (for bills only), rationale for restrictions, health outcomes cited in justification, and use of evidence were captured from waiver requests and bills. Related to waiver implementation, items related to the planned implementation supports described, planned communications to SNAP participants and retailers, anticipated impacts on state agency staff, state agency budgets and retailers were captured from the waiver requests as more detail was provided in these documents than in the approvals. Final food and beverage groups restricted and their definitions, as well as final evaluation plans, were captured from the waiver approvals. Some items in the codebook were open-ended (e.g., copy/paste exact definition of food or beverage), and others had pre-specified response options (e.g., planned communication strategies).

### Coding

After the initial codebook was developed, it was piloted by all authors involved in coding (EWD, ED, MAL) on a random sample of one document of each type, and then the codebook was refined to improve clarity and consistency. Each document was then double-coded, and discrepancies were resolved by consensus. One request and an approval, from Montana, was single-coded by EWD because this state received approval after data collection was completed but prior to manuscript submission. All initial coding was conducted in Qualtrics.

### Analysis

Descriptive statistics on reconciled coded content were conducted in Microsoft Excel (version 16.106.3). Existing information on state-level party affiliation was merged with legislative data to provide further contextual information about the waiver adoption process^13,14^. For comparisons of foods or beverages restricted when definitions were considered, commonly accepted definitions for common beverage types (e.g., fruit drinks, energy drinks, soda) from What We Eat in America^15^ were used by one team member (EWD) to determine which beverage categories would likely be restricted based on the state’s definition. This study was deemed exempt, non-human subjects research by Viable Insights Review Board (VIRB) for the Protection of Human Subjects in Research (REF000245).

## Results

### State Bills

The final sample of state bills included 71 bills that were introduced in 35 states from 2025 through March of 2026 (Figure 1). Only seven (10%) of the bills were introduced by a bipartisan group of lawmakers, with the remainder being introduced by Republican lawmakers. Slightly more than half (n=42, 59%) of bills were introduced in states with a Republican Governor, Republican-controlled House, and Republican-controlled Senate (i.e., Republican trifecta), and 14 (20%) were introduced in states with a Democratic trifecta. Three bills (4%) proposed both restrictions and additions. Bills mostly proposed restricting candy (n=46, 65%) and/or soft drinks (n=42, 59%) (Figure 2).

**Figure 1:**
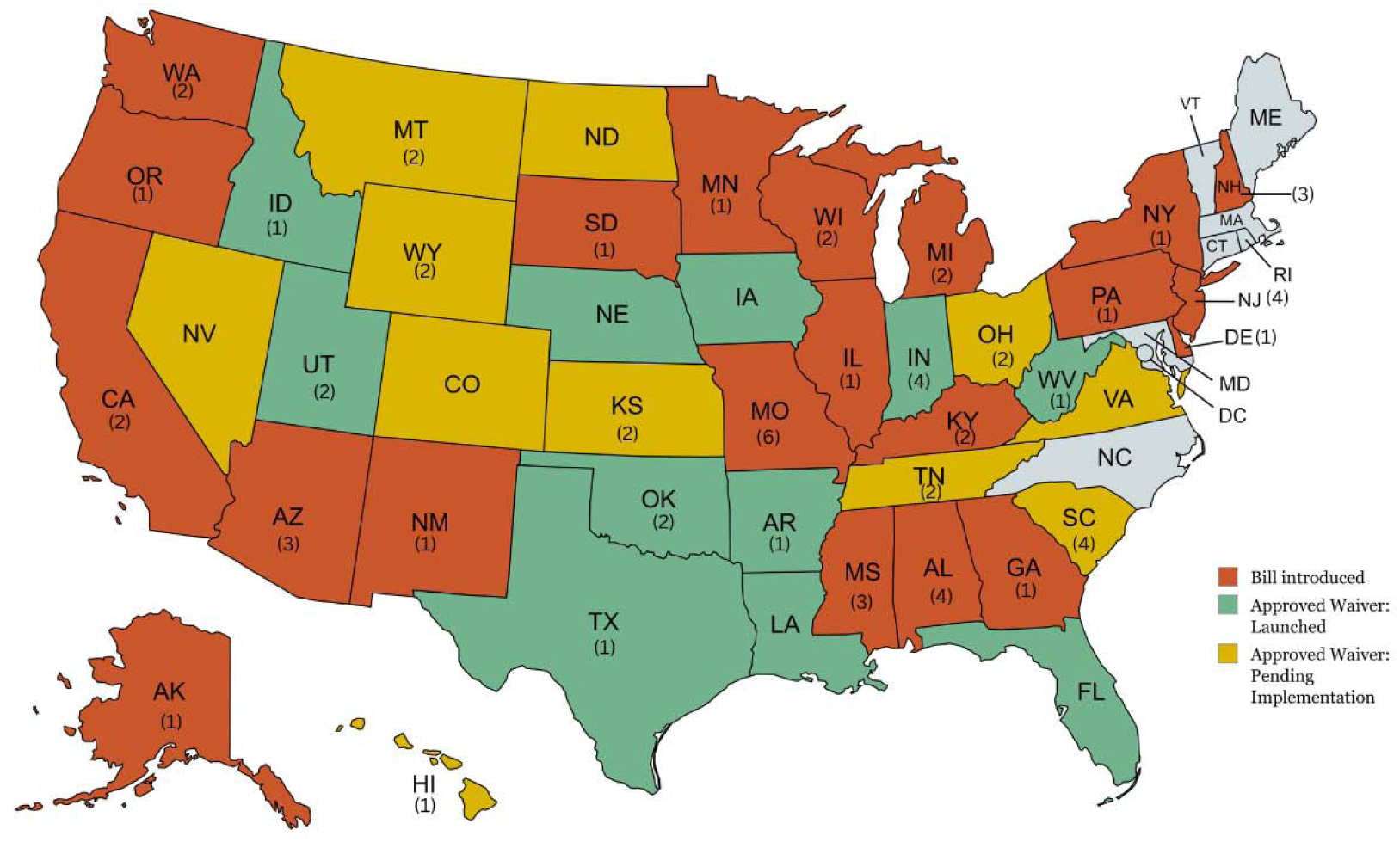
States that have introduced Supplemental Nutrition Assistance Program (SNAP) food restriction waiver-related legislation, received approval to implement a food restriction waiver, or are currently implementing a food restriction waiver as of June 2026 Numbers in parentheses are the number of bills introduced during the study period

**Figure 2:**
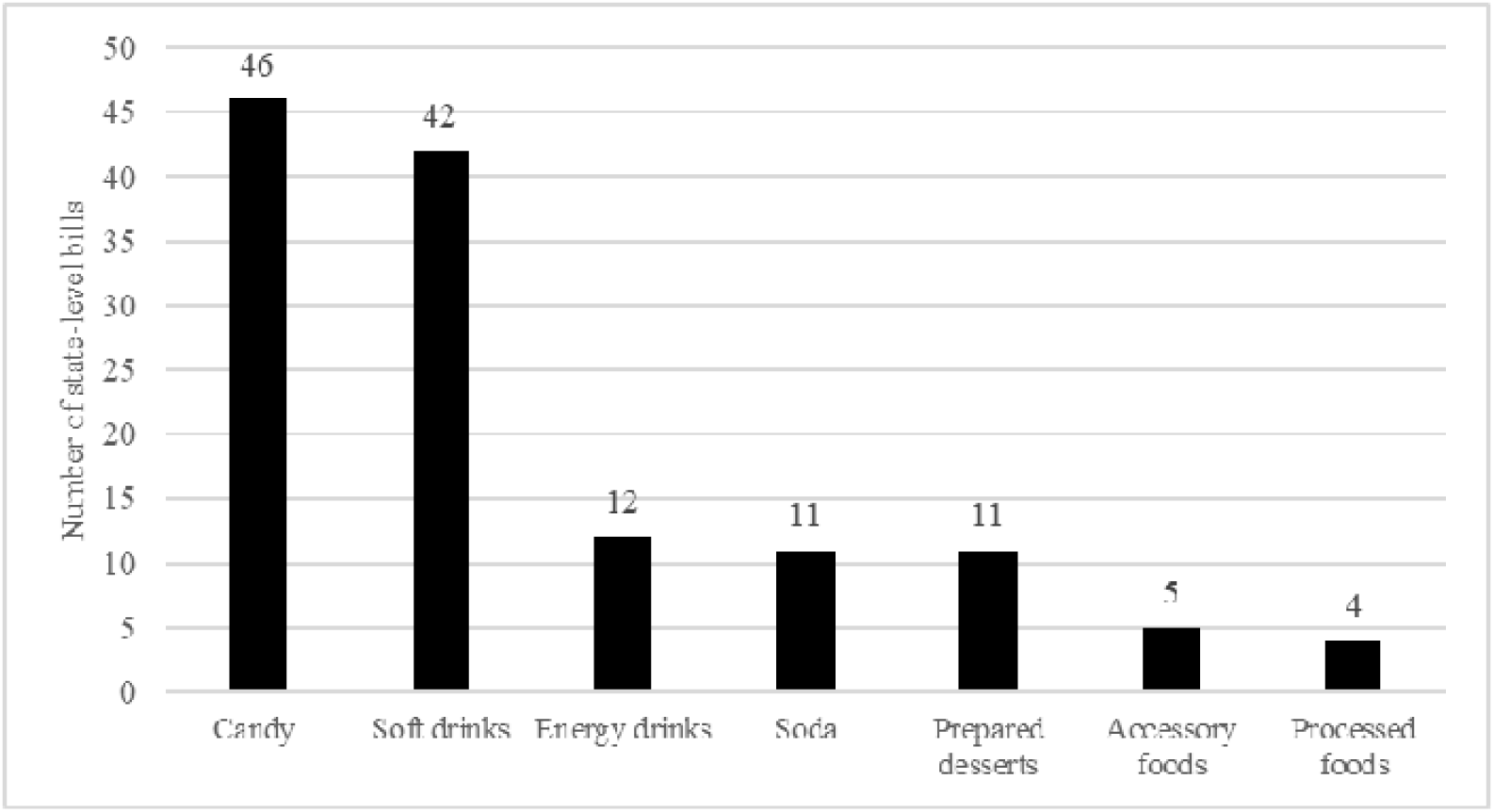
Food and beverage categories included in proposed Supplemental Nutrition Assistance Program (SNAP) food restriction waiver-related state bills from Jan 2025-March 2026 (n=71)

### Framing of Bills

Most bills did not include a justification for pursuing the SNAP food restriction waiver (n=55, 77%), but among those that did, justifications included improving diet and/or health outcomes (n=12, 17%), aligning SNAP with its intended purpose (n=10, 14%), increasing cost effectiveness (n=9, 13%), or aligning SNAP with other federal nutrition assistance programs that have nutrition standards (n=5, 7%) (Table 1). Most bills did not cite specific health outcomes of concern as motivation for pursuing the waiver (n=58, 82%); however, among those that did, 10 (14%) cited poor dietary intake, and eight (11%) cited obesity.

**Table 1:**
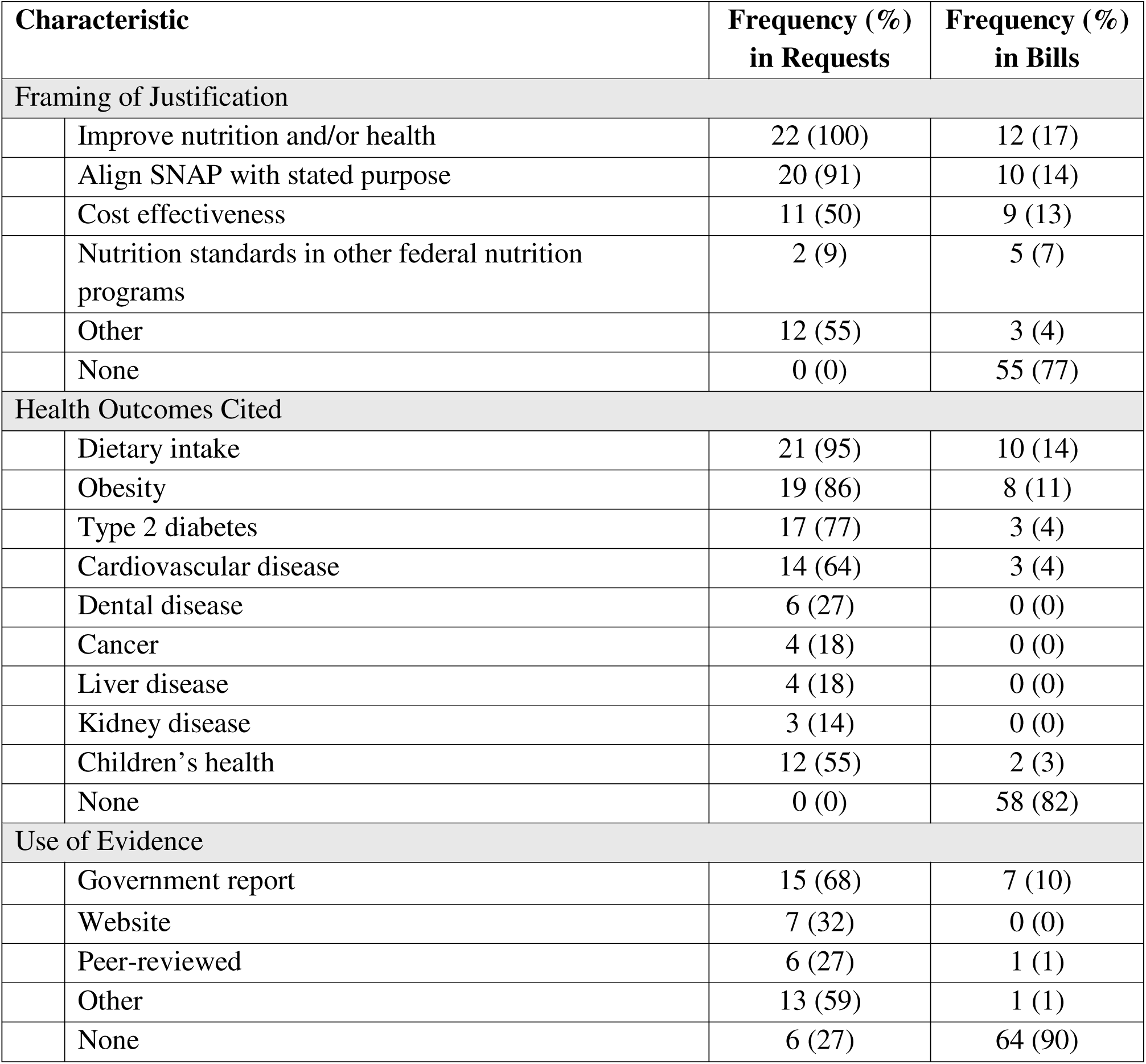
Justification framing, health outcomes cited, and use of evidence in Supplemental Nutrition Assistance Program (SNAP) food restriction waiver requests (n=22) and state bills (n=71)

| Characteristic |  | Frequency (%)<br>in Requests | Frequency (%)<br>in Bills |
| --- | --- | --- | --- |
| Framing of Justification |  |  |  |
|  | Improve nutrition and/or health | 22 (100) | 12 (17) |
|  | Align SNAP with stated purpose | 20 (91) | 10 (14) |
|  | Cost effectiveness | 11 (50) | 9 (13) |
|  | Nutrition standards in other federal nutrition programs | 2 (9) | 5 (7) |
|  | Other | 12 (55) | 3 (4) |
|  | None | 0 (0) | 55 (77) |
| Health Outcomes Cited |  |  |  |
|  | Dietary intake | 21 (95) | 10 (14) |
|  | Obesity | 19 (86) | 8 (11) |
|  | Type 2 diabetes | 17 (77) | 3 (4) |
|  | Cardiovascular disease | 14 (64) | 3 (4) |
|  | Dental disease | 6 (27) | 0 (0) |
|  | Cancer | 4 (18) | 0 (0) |
|  | Liver disease | 4 (18) | 0 (0) |
|  | Kidney disease | 3 (14) | 0 (0) |
|  | Children's health | 12 (55) | 2 (3) |
|  | None | 0 (0) | 58 (82) |
| Use of Evidence |  |  |  |
|  | Government report | 15 (68) | 7 (10) |
|  | Website | 7 (32) | 0 (0) |
|  | Peer-reviewed | 6 (27) | 1 (1) |
|  | Other | 13 (59) | 1 (1) |
|  | None | 6 (27) | 64 (90) |

### Waiver Requests and Approvals

The final sample included 22 waiver requests and 23 waiver approvals. One waiver request was not publicly available on the USDA website and, therefore, was not included. All approved waiver requests restrict the use of SNAP benefits to purchase specific types of beverages, and 15 (65%) waivers restrict the use of SNAP benefits to purchase specific types of food. Using the terms described in the approved waivers, the restricted beverages were soda or soft drinks (n=14, 61%), sweetened or high-sugar beverages (n=9, 39%), energy drinks (n=7, 23%), unhealthy drinks (n=2, 9%), fruit drinks (n=1, 4%), and diet drinks (n=1, 4%). The restricted foods included candy (n=15, 65%), prepared desserts (n=3, 13%), and processed foods (n=1, 4%).

### Framing of Request

In terms of the framing of the justification for the waiver requests to USDA, all states (22) mentioned improving diet or health outcomes, 20 (91%) cited aligning SNAP with its intended purpose (which varied among states), 11 (50%) states cited cost effectiveness, two (9%) mentioned aligning SNAP with other federal nutrition assistance programs with nutrition standards, and eight (36%) mentioned other justifications such as aligning with the Dietary Guidelines or supporting local agriculture (Table 1). Health outcomes commonly included in the justification were low-quality dietary intake (n=21, 95%), obesity (n=19, 86%), type 2 diabetes (n=17, 77%), and cardiovascular disease (n=14, 64%) (Table 1). Twelve states cited children’s health in their waiver request justification. Most states (n=16, 73%) cited some type of evidence in their waiver request. Predominantly, states cited government reports (n=15, 68%), followed by websites (n=7, 32%), then peer-reviewed sources (n=6, 27%) (Table 1).

### Planned Implementation and Communication Strategies

For implementation supports described in their requests, 19 (86%) states described participant communications plans, 18 (82%) described retailer communications plans or education, 15 (68%) described nutrition education, 11 (50%) described staff training, and 10 (45%) described collaborations with broader community groups. The planned SNAP participant and retailer communications strategies (e.g., website content, emails,) that states described in their requests are summarized in Table 3. Three states (14%) described piloting some component of the SNAP food restriction waiver prior to full implementation, such as starting with one food group or a subset of retailers. Fourteen states (64%) mentioned collaborating with SNAP-Education (SNAP-Ed) in their waiver requests. Thirteen of these states stated SNAP-Ed would do nutrition education, and five said SNAP-Ed would be engaged in the evaluation of the food restriction waiver. While some states (n=13, 57%) stated in their request that they anticipated being able to use existing staff and resources for implementation, others described anticipated increases in costs for staffing, technical assistance, and evaluation, or that costs would be borne by retailers.

### Food and Beverage Category Definitions

States varied considerably in how they defined restricted beverage categories such as soft drinks, soda, and sweetened beverages. Seventeen states (74%) included artificially sweetened beverages in their definitions of restricted beverages. Ten states (43%) limited restrictions to carbonated beverages with sweeteners, while 10 (43%) states applied restrictions to any non-alcoholic sweetened beverage regardless of carbonation (Table 2). When these broader definitions were considered, a wider variety of beverages could be restricted than what is captured using the terms in the waiver alone. For example, while seven states (30%) explicitly mentioned restricting energy drinks in their waivers, 21 (95%) states could restrict energy drinks based on their definitions of restricted beverages such as soda or soft drinks (e.g., any non-alcoholic beverage that contains natural or artificial sweeteners) (Figure 3). Seven states (32%) had quantitative limits on the amount of sweetener in a beverage as part of their definitions (e.g., five grams), though the serving size for the quantitative limit (grams per ounce) was only specified in one of the definitions. Two states (9%) also used ingredient order in their definitions, restricting beverages with sweeteners as one of the first two ingredients (Table 2). For candy, some states used the definition of candy often found in the sales tax code (i.e., preparation of sugar, honey, or other natural or artificial sweeteners in combination with chocolate, fruits, nuts or other ingredients or flavorings in the form of bars, drops, or pieces)^16^. A small number of states explicitly stated that candy requiring refrigeration (n=3, 20% of states restricting candy) or containing flour (n=2, 13% of states restricting candy) would not be restricted.

**Figure 3:**
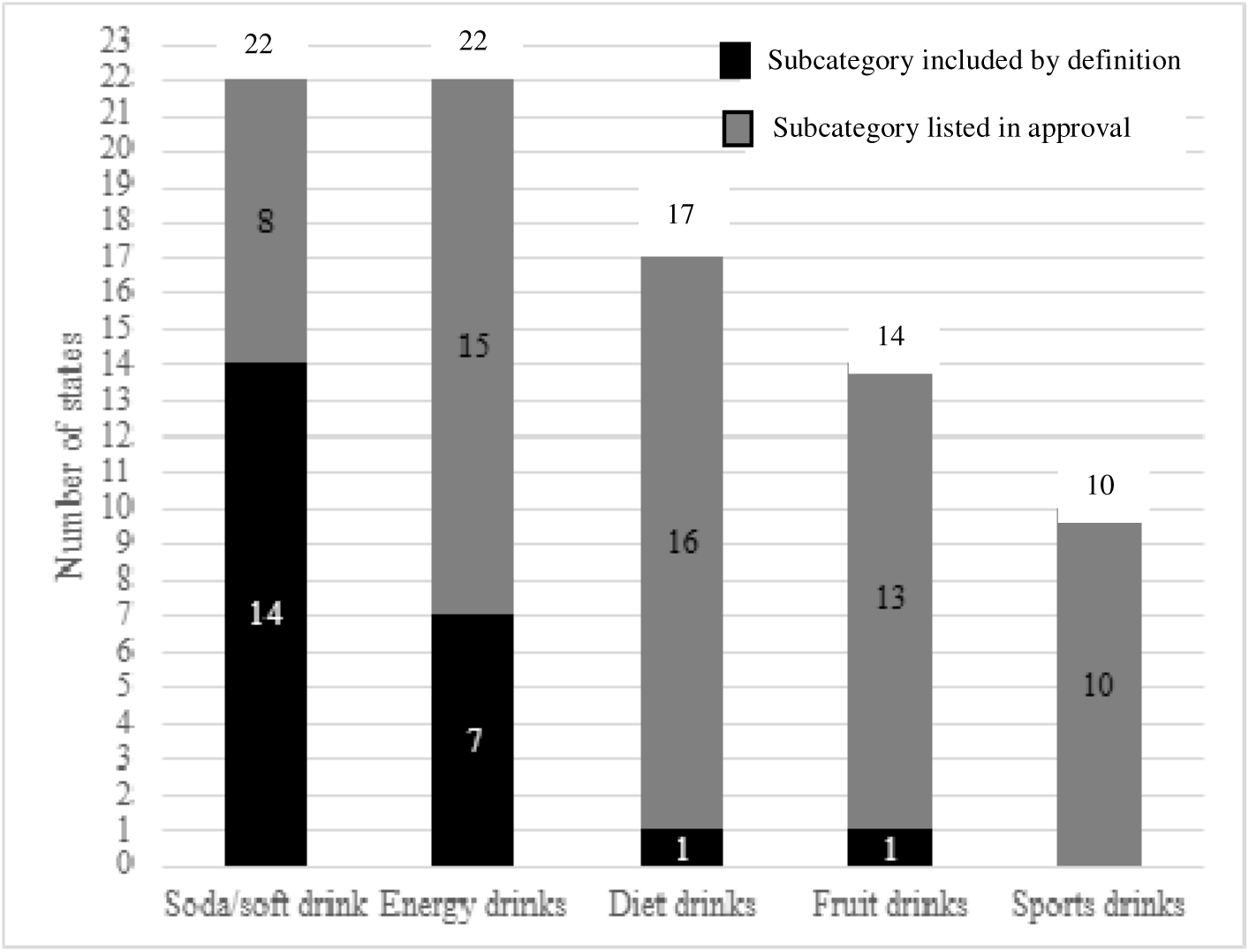
Subcategories of beverages restricted when using food or beverage categories named in approvals compared to the broader scope of beverages likely restricted when considering state-level definitions of categories from waiver approvals^a^ (n=22^b^) ^a^Comparisons are based on definitions provided in waiver approvals as of June 2026 and using commonly accepted definitions of beverage subcategories from NHANES WWEIA ^b^For energy drinks and sports drinks, one state did not provide sufficient information in the waiver approval to make a determination, so n=22 for those subcategories

**Table 2.**
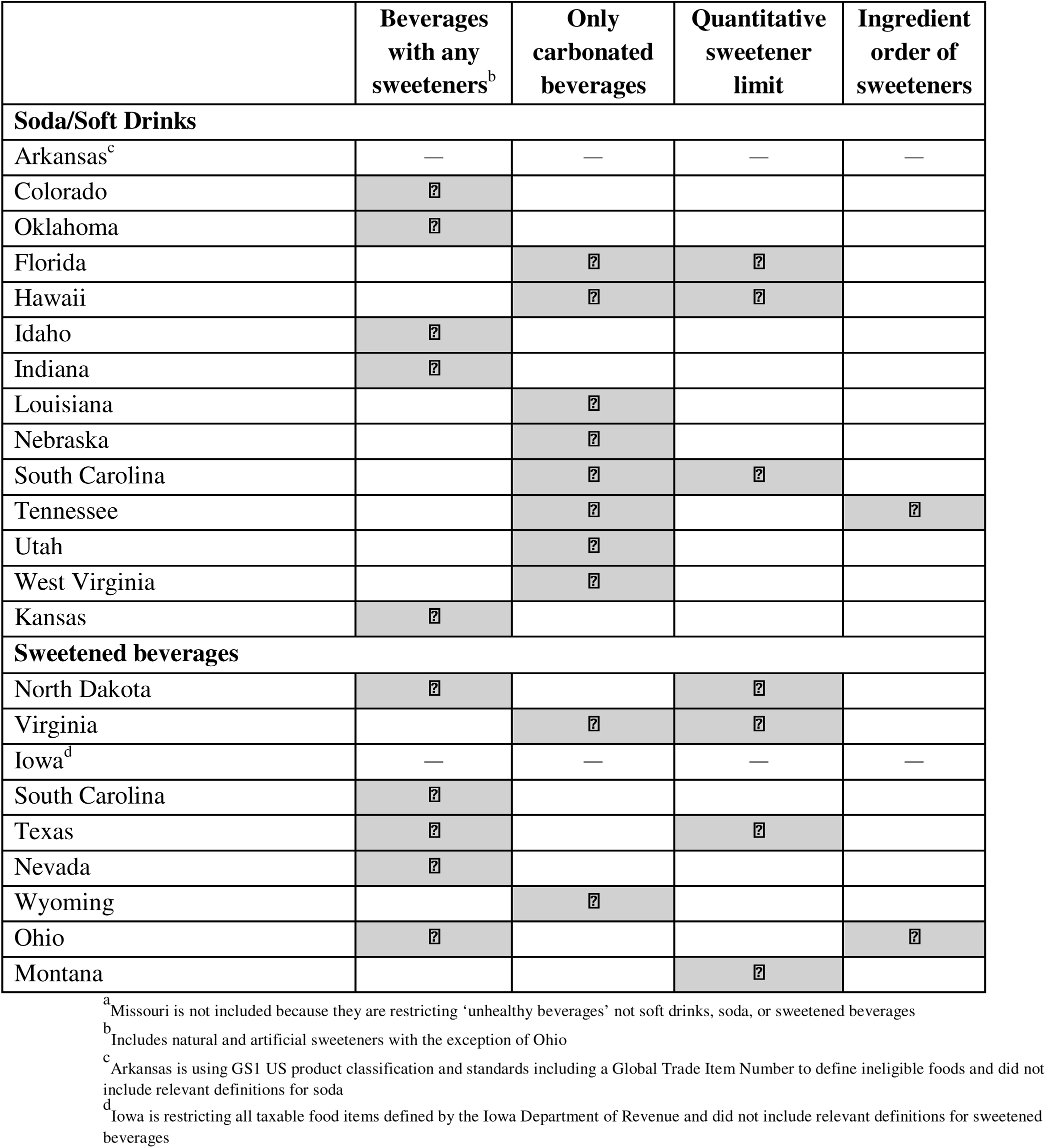
Key characteristics of definitions of soda/soft drinks, and sweetened beverages from approved waivers at the state-level (n=22 states^a^)

|  | **Beverages**<br>**with any**<br>**sweeteners<sup>b</sup>** | **Only**<br>**carbonated**<br>**beverages** | **Quantitative**<br>**sweetener**<br>**limit** | **Ingredient**<br>**order of**<br>**sweeteners** |
| --- | --- | --- | --- | --- |
| **Soda/Soft Drinks** |  |  |  |  |
| Arkansas<sup>c</sup> | — | — | — | — |
| Colorado | ? |  |  |  |
| Oklahoma | ? |  |  |  |
| Florida |  | ? | ? |  |
| Hawaii |  | ? | ? |  |
| Idaho | ? |  |  |  |
| Indiana | ? |  |  |  |
| Louisiana |  | ? |  |  |
| Nebraska |  | ? |  |  |
| South Carolina |  | ? | ? |  |
| Tennessee |  | ? |  | ? |
| Utah |  | ? |  |  |
| West Virginia |  | ? |  |  |
| Kansas | ? |  |  |  |
| **Sweetened beverages** |  |  |  |  |
| North Dakota | ? |  | ? |  |
| Virginia |  | ? | ? |  |
| Iowa<sup>d</sup> | — | — | — | — |
| South Carolina | ? |  |  |  |
| Texas | ? |  | ? |  |
| Nevada | ? |  |  |  |
| Wyoming |  | ? |  |  |
| Ohio | ? |  |  | ? |
| Montana |  |  | ? |  |

<sup>a</sup> Missouri is not included because they are restricting ‘unhealthy beverages’ not soft drinks, soda, or sweetened beverages
<sup>b</sup> Includes natural and artificial sweeteners with the exception of Ohio
<sup>c</sup> Arkansas is using GS1 US product classification and standards including a Global Trade Item Number to define ineligible foods and did not include relevant definitions for soda
<sup>d</sup> Iowa is restricting all taxable food items defined by the Iowa Department of Revenue and did not include relevant definitions for sweetened beverages

### Evaluation Plans

The evaluations of food restriction waivers in all approved states (n=23) require data to be collected on: meals eaten away from home; purchases of ‘unhealthy’ foods not restricted by the food restriction waiver; payment substitution for restricted items; SNAP participant awareness of restricted foods; impacts on SNAP participants’ shopping routines; and participant, retailer, advocacy, community, and retailer group satisfaction (Table 3). Some states proposed additional evaluation outcomes such as cross-border shopping (n=19, 83%) and effectiveness of communication efforts (n=8, 35%). The data collection methods described varied considerably, with all states proposing to use SNAP participant surveys, SNAP redemption data, transaction data, and complaint data (Table 3). About half (n=11, 48%) of the states with approved waivers mentioned using retailer surveys, six (26%) mentioned 24-hr recalls or food records, and five (22%) mentioned existing administrative or state-level datasets, including Medicaid data.

**Table 3:** States’ planned food restriction waiver-related communication strategies for Supplemental Nutrition Assistance Program (SNAP) participants and SNAP-authorized retailers included in waiver requests (n=22) and planned evaluation data collection methods and outcomes included in waiver approvals (n=23)

| Characteristic |  | Frequency (%) |
| --- | --- | --- |
| SNAP participant communications |  |  |
|  | Direct communications (email, phone, mail) | 14(64) |
|  | Website | 14 (64) |
|  | Staff talking points | 12 (55) |
|  | Social media | 10 (45) |
|  | Community partner collaboration | 9 (41) |
|  | Press release | 8(36) |
|  | Other | 13(59) |
|  | No specific strategies mentioned | 4(18) |
| Retailer communications |  |  |
|  | Signage | 11 (50) |
|  | Direct communication | 10 (45) |
|  | Website | 9(41) |
|  | FAQ | 7(32) |
|  | Staff talking points | 5(23) |
|  | Social media | 2(9) |
|  | Webinar | 2(9) |
|  | No specific strategies mentioned | 5 (23) |
| Data collection method |  |  |
|  | Existing compliment and complaint tracking | 23(100) |
|  | Retailer transaction data | 23(100) |
|  | SNAP participant surveys | 23(100) |
|  | SNAP redemption data | 23 (100) |
|  | Retailer surveys | 11(48) |
|  | Other secondary data sources | 5 (22) |
|  | Medicaid claims | 2(9) |
| Evaluation outcome |  |  |
|  | Advocacy and community group satisfaction | 23(100) |
|  | Changes in shopping routines | 23(100) |
|  | Meals away from home | 23(100) |
|  | Non-SNAP payment substitution for restricted items | 23(100) |
|  | Participant knowledge and awareness | 23(100) |
|  | Participant satisfaction | 23(100) |
|  | Purchases of unhealthy, unrestricted foods | 23(100) |
|  | Retailer satisfaction | 23(100) |
|  | Cross-border shopping | 19(83) |
|  | Effectiveness of communications | 8(35) |

## Discussion

Findings from this content analysis suggest the landscape of state-level SNAP food restriction waivers remains highly variable, and the volume of bills introduced since 2025 indicates food restriction waivers are a policy priority for many states. The present study describes key characteristics related to the adoption and implementation of early adopting states that other states interested in adopting SNAP food restriction waivers may use to inform their efforts, enhance the overall effectiveness of waivers on intended outcomes, and minimize unintended consequences. The existing state-level variability in waiver elements such as foods and beverages included, implementation supports, and communication strategies will likely have substantive impacts on the experiences of key groups such as SNAP participants, state agency staff, and retailers. The present study builds on prior similar work by others^7,17^ by including a focus on content related to waiver implementation, an updated sample of states with approved waivers prior to the June 2026 court ruling, and a review of recent state bills related to food restriction waivers.

There is a high degree of variability and complexity across states in the food and beverage categories restricted and definitions of categories. The variability across states has important implications for retailers and participants. SNAP must remain interoperable, meaning benefits must work across states, so participants shopping in different states will need to adapt to different definitions, and retailers operating across multiple states must invest resources in implementing different restrictions. In terms of complexity, many of the definitions, such as soda being defined as any non-alcoholic beverage with natural or artificial sweeteners, will require SNAP participants and retailers to perform time-consuming tasks that require a high level of nutrition literacy, such as reviewing nutrition labels and ingredients lists to identify caloric and non-caloric sweeteners or stimulants to properly identify restricted foods or beverages. Additionally, some definitions as currently written may unintentionally restrict health-promoting beverages; however, definitions are subject to change, and as of the start of July 2026, nine of the 23 states have already received approval to modify their original definitions^3^.

State agencies are making concerted efforts to operationalize complex definitions and communicate restricted beverage categories to SNAP participants and retailers; however, as has been well documented in the Special Supplemental Nutrition Program for Women, Infants, and Children (WIC) literature, participants may experience challenging or stigmatizing experiences in the retail environment trying to avoid restricted foods which may inadvertently influence willingness to participate in SNAP over the long term^18–20^. Many restricted category definitions are broad and based on existing tax codes with the intent of facilitating retailer implementation, and a reasonable public health argument could be made for restricting all beverages with caloric sweeteners if the end goal of these waivers is to reduce total added sugar consumption. However, the tradeoffs between retailer and SNAP participant experiences must be weighed carefully in defining restricted categories as future states adopt similar waivers or if the US moves toward federal-level restrictions. On the retail side of operationalizing complex definitions, some, but not all states are providing access to a list of restricted products at the Universal Product Code (UPC)-level^21^, but it is not known whether this sort of resource will be available or updated as time goes on and, without adequate technical assistance, retailers may fall out of compliance or decide to no longer be a SNAP-authorized retailer. If SNAP food restriction waivers continue to be adopted and are maintained over the long-term, investments in infrastructure similar to what has been done in WIC, such as maintaining Approved/Authorized Product Lists (APLs) and participant-facing applications, (e.g., Arkansas’s SNAP Shopper App that identifies approved foods in the retail environment), may become necessary to support retailer and household-level SNAP participation.

States with approved waivers describe a variety of planned implementation supports for participants and retailers, including comprehensive communication plans required by USDA. One way of supporting implementation would be to test implementation on a smaller scale initially; however, only three states with approved waivers mentioned piloting the food restriction waiver on a small scale before full implementation. A second strategy for supporting implementation could be providing longer than six months between enactment and implementation^9^; however, many states with approved waivers had abbreviated timelines between approval and implementation. Third, providing adequate resources and staffing are known implementation facilitators, but many states explicitly stated they would maintain cost neutrality and use existing staff and agency resources for waiver implementation, while more recent requests included explicit funding requests for more staff. The high volume of co-occurring SNAP policy changes may strain the existing social service agency workforce, so understanding state agency staff members’ experiences and downstream effects of staff capacity on implementation in states with varying approaches to staffing may be important. Fourth, it was originally expected that states would leverage SNAP-Ed for implementation support. More than half of states with approved waivers stated SNAP-Ed partners would provide either nutrition education programming related to the SNAP food restriction waiver to participants or evaluation support. Elimination of SNAP-Ed federal funding as of September 2026 presents another important area for future inquiry and a potential gap for academic and community partners to fill in terms of education and evaluation support. Finally, while presence of state or local nutrition incentive programs is not systematically included in the documents reviewed for the present study, an increased focus on healthy food incentives may be warranted if the goal of SNAP food restrictions is to improve participants’ dietary intakes and health^22–24^. Overall, future studies of state-level implementation of the SNAP food restriction waivers and understanding contextual factors that support or hinder implementation will be valuable.

In waiver requests, states laid out comprehensive communication plans for SNAP-participating households and retailers, which is another factor commonly cited in determinant frameworks as a critical component for implementation^25,26^. Ongoing monitoring and documentation of actual state-level communications and narratives about waivers in the public discourse will provide important contextual information about SNAP participant and retailer awareness and attitudes and how that may interact with the impact of waivers on nutrition and participation-related outcomes. Additionally, monitoring potential differential impacts of implemented communication strategies on awareness and downstream outcomes such as SNAP participation among subgroups of SNAP participants or retailers may be important, as factors such as primary language and digital or health literacy will likely influence the effectiveness of communication strategies^27–29^.

Findings from the review of state bills highlighted that most US states have introduced bills related to SNAP food restriction waivers, and proposed restrictions were variable in terms of food and beverage categories but predominantly focused on candy and soft drinks. The results presented here can be used as a tool for researchers and practitioners interested in planning programmatic efforts to support implementation and impact evaluations related to future waivers. Additionally, as described, later adopting states can use this information to inform and improve on waiver design and implementation strategies. Documentation of the framing of proposed bills and waiver requests also provides useful contextual information about waiver adoption that could be used to inform work in similar policy areas such as SNAP incentives or policymaker-focused dissemination research^30^. Development of more robust SNAP policy surveillance systems would be worthwhile^31^, as the work presented here was intended to provide researchers, practitioners, and other relevant groups with a sense of trends in current SNAP food restriction waiver bills, not as an exhaustive tool used for evaluation purposes.

Strengths of the current study include providing a comprehensive review of the contents of waiver requests and approvals, which will be highly informative for future state-level or multi-state evaluations. Additionally, the inclusion of state bills is a novel addition that can serve as a planning tool for researchers and practitioners. Additionally, the data presented here in terms of implementation and communications are what states have planned, not what states may implement, highlighting the need for ongoing studies of implementation.

The current state of SNAP food restriction waivers is highly variable and shifting rapidly. Surveillance and documentation of key elements of early adopters of SNAP food restriction waivers will be a critical resource for researchers and practitioners studying the process of implementation and the intended and unintended impacts of waivers on SNAP participants, retailers, and other key groups. Future research will be needed as additional states adopt SNAP food restriction waivers to understand ongoing implementation, impacts on SNAP participants’ experiences, diet and health, and what may be needed for successful maintenance of waivers from diverse groups’ perspectives.

## Data Availability

All data produced in the present study are available upon reasonable request to the authors

## Notes

### Competing Interest Statement

A.J.M. has received consultancy fees from non-profit advocacy organizations, academic institutions, philanthropies and government agencies for work related to public health nutrition, including SNAP.

### Funding

- Healthy Eating Resarch
- Bloomberg Philanthropies: 2025-153247

