## Supplemental File 1 for "State-Level Variability in Implementation Approaches to SNAP Food Restriction Waivers: A Content Analysis of Bills, Waiver Requests, and Waiver Approvals"

**Supplemental File 1:** Items and response options for the codebooks for Supplemental Nutrition Assistance Program (SNAP) food restriction waiver requests, waiver approvals, and bills that was used to program each codebook into Qualtrics

| **Item** | **Response options** |
| --- | --- |
| **Document Information** | |
| State | Drop down list of all states |
| Team Member | List of team members |
| Document type | State senate bill  State house bill  Other state legislation  Waiver request  Waiver approval |
| **Waiver Request Items** | |
| Date submitted (mm/dd/yyyy) | Text box |
| Requesting official name | Text box |
| Requesting official title | Text box |
| Requesting official email | Text box |
| Requesting official phone number | Text box |
| State SNAP contact name | Text box |
| State SNAP contact title | Text box |
| State SNAP contact phone number | Text box |
| State SNAP contact email | Text box |
| Regional SNAP contact name | Text box |
| Regional SNAP contact phone number | Text box |
| Regional SNAP contact email | Text box |
| Other names/contact information listed on request | Text box |
| New foods or beverages restricted (in their words-exact definition captured later) [select all that apply]  For example, if they say they are restricting "Soda" but their definition includes artificially sweetened sodas, only select Soda do not select Diet Soda. | Soda  Soft drinks  Sweetened drinks  Sweetened water  Diet soda/low/no calorie soda  Fruit and vegetable drinks  Unhealthy drinks  Sports drinks  Energy drinks  Candy  Prepared desserts  Seeds for food producing  Food producing plants  Fruit leather  Sweetened baking chocolate  Candy or chocolate coated fruit  Granola bars  Sweetened coconut  Marshmallows  Trail mix  Caramel or kettle corn  Other:______ |
| Definition(s) of restricted foods/beverages | Text box |
| Definition(s) of beverages NOT included in restrictions | Text box |
| Rationale for restriction includes [select all that apply] | Cost effectiveness/cost savings (including health care costs)  To align with the stated purpose/goals of SNAP  Diet and/or health outcomes  Existing restrictions in other federal nutrition assistance programs (e.g., WIC, school meals)  Increased SNAP household purchasing power  Other: _____ |
| Stated intent/purpose of SNAP (not restrictions, the whole program) | Text box |
| Health outcomes cited in justification for restrictions [select all that apply] | Low diet quality/poor dietary intake  Obesity  Type 2 diabetes  Cardiovascular disease  Cancer  Kidney disease  Dental disease  Liver disease  Other:______ |
| Mentions children’s health in justification for restrictions | Yes  No  Don’t know/can’t tell |
| Children’s health outcomes cited [select all that apply] | Low diet quality/poor dietary intake  Obesity  Type 2 diabetes  Cardiovascular disease  Cancer  Kidney disease  Dental disease  Liver disease  Not applicable/none stated  Other:______ |
| Types of literature cited [select all that apply] | Government report(s) or link to government report(s)  Peer-reviewed publication  Other report  Website  None  Other: _______ |
| Links to literature cited | Text box |
| Mentions “Make America Healthy Again” or making [state name] healthy again | Yes  No |
| Anticipated implementation date | Text box |
| Do they mention they will pilot these restrictions on a smaller scale before fully implementing them? | Yes  No |
| If yes, what are the details of the pilot plans | Text box |
| Implementation supports described [select all that apply] | Collaboration with other SAs  Collaboration with other community groups  SNAP participant nutrition education about healthy food/beverage options  SNAP participant communications/communication plan related to the waiver  Retailer discussions/work group  Retailer education  Retailer communications/communication plan  SNAP staff training  Other: ________ |
| Timeline listed for communication plan | Text box |
| Intended audience for communication plan | SNAP participating households  General public  Advocates  Retailers  Not stated  No communication plan(s) described  Other:_____ |
| SNAP participant communication strategies about waiver described | Website content  Press release  Community partner collaboration  Brochures  Mail  Social media  Phone calls  Emails  Text messages  In-person programs  Infographics  Talking points for SNAP staff or communication training  None  Other:_______ |
| Retailer communication strategies about waiver described [select all that apply] | Website content  Social media  Infographics  Signage or posters  Emails  Mailers  Webinars  Talking points for retailers or communication training  FAQ document  None  Other:_______ |
| SNAP Education mentioned | Yes  No |
| If yes, what role is described for SNAP Ed | Text box |
| Anticipated impacts/outcomes of waiver implementation described in anticipated outcomes, impacts on state agency, or similar section (s) [select all that apply] | Improvements in diet  Improvements in health outcomes  Decreases in food access  No impact on SA staff time  No impact on SA budget  No impact on retailer expenses  None mentioned  Other: ______ |
| Retail trade group mentioned in request | Yes/No |
| Retail trade group contact information |  |
| Restrictions apply to summer EBT/pEBT | Yes  No  Not stated |
| Length of time retailers have to implement | Text box |
| Compliance plan for retailers described | Attestation statement/self-report  Existing reporting channels  Credible tips from the public  Secret shoppers  Monitoring program  Tracking complaints  None mentioned  Other:______ |
| Evaluation outcomes planned. | Obesity/BMI  Type 2 diabetes  Hypertension  CVD  Economic impacts  Food consumption  Food purchases  Self-reported meals away from home  Self-reported payment substitution  Emotional impacts  Participant waiver awareness  Stigma  Food access  SNAP participation  Retailer awareness  Retailer technical capabilities  Retailer implementation costs  Retailer barriers to adoption  SNAP redemption  Retailer SNAP participation rates  Geographic retailer coverage  Out-of-state SNAP redemption  Other:______ |
| Evaluation data sources | Medicaid claims data  WIC data  Other existing state-level datasets/surveillance systems  Participant surveys  Retailer readiness and needs assessment  Retailer surveys  Retailer data (transaction data-not EBT redemption data)  Listening sessions with retailers  EBT redemption data  Participant complaints  Retailer complaints |
| Evaluation partner or potential partner specified | Yes  No |
| If yes, list partner | Text box |
| Other notes/important information not captured above | Text box |
| Waiver Approval Items | |
| Waiver start | Month/Day/Year |
| Waiver duration | Text box |
| Extension possible | Yes  No |
| If yes, duration of possible extensions | Text box |
| New foods or beverages restricted [select all that apply] | Soda  Soft drinks  Sweetened drinks  Sweetened water  Diet soda/low/no calorie soda  Fruit and vegetable drinks  Unhealthy drinks  Sports drinks  Energy drinks  Candy  Prepared desserts  Seeds for food producing  Food producing plants  Fruit leather  Sweetened baking chocolate  Candy or chocolate coated fruit  Granola bars  Sweetened coconut  Marshmallows  Trail mix  Caramel or kettle corn |
| Definition(s) of restricted foods/beverages | Text box |
| Definition(s) of beverages NOT included in restrictions | Text box |
| Required communications to retailers | Communications plan  Website  Centralized communication resource  Other:______ |
| Required communications to SNAP participating households | Communication plan  Website  Other:______ |
| Evaluation plan required | Yes  No |
| If yes, required and proposed evaluation outcomes | Meals and food eaten outside the home/food not purchased at SNAP-authorized retailers  Purchases of unhealthy foods not restricted  Non-SNAP dollars spent on restricted items  Cross-border shopping  SNAP client confidence and ability to identify restricted items  SNAP participant awareness  Impact on participants’ shopping routines (e.g., distance to store, frequency of trips)  SNAP client compliments, complaints, or hearings  Retailer compliments or complaints  Effectiveness of communication and implementation efforts |
| If yes, required and proposed evaluation data collection methods  If there are more extensive evaluation methods proposed beyond basic requirements, copy/paste those in "Other" | Complaint tracking and satisfaction data  Retailer surveys and interviews  SNAP participant surveys  Retailer transaction data  Redemption data  Medicaid claims  Other existing state-level secondary data  Other_______ |
| Evaluation partner or potential partner named | Yes  No |
| Name of evaluation partner |  |
| Retailer compliance and monitoring plan required | Yes  No |
| Other notes/important information not captured above | Text box |
| **Bill Items** | |
| Date introduced | Month/day/year |
| Number of sponsors | Dropdown 1-10 |
| Bill sponsor name | Text box |
| Bill sponsor party | Democrat  Green Party  Independent  Libertarian  No affiliation  Progressive  Republican  N/A  Other |
| Bill status | Passed  Failed  Introduced  Sent to Governor  Other: ______ |
| Does the bill include suggested SNAP restrictions, additions, or both? | Restrictions  Additions  Both |
| New foods or beverages restricted [select all that apply] | Soda  Soft drinks  Sugar sweetened beverage  Sweetened water  Diet soda  Candy  Seeds for food producing  Food producing plants  Fruit leather  Prepared desserts  Processed foods  Sweetened baking chocolate  Candy or chocolate coated fruit  Granola bars  Sweetened coconut  Marshmallows  Energy drinks  Sports drinks  Trail mix  Caramel or kettle corn  Unhealthy drinks  Other: |
| Definition(s) of restricted foods/beverages | Text box |
| Definition(s) of beverages NOT included in restrictions | Text box |
| New foods or beverages added (in their words-exact definition captured later) [select all that apply] | Rotisserie/hot prepared chicken  Other: |
| Definitions of added foods/beverages (copy/paste exact words from request for any food or beverage selected) | Text box |
| Rationale for restriction includes [select all that apply] | Cost effectiveness/cost savings (including health care costs)  Original intent of SNAP  Health outcomes  Existing restrictions in other federal nutrition assistance programs (e.g., WIC, school meals)  Increased SNAP household purchasing power  Other: _____ |
| Stated intent/purpose of SNAP (not restrictions, the whole program) | Text box |
| Health outcomes cited in justification for restrictions [select all that apply] | Low diet quality/poor dietary intake  Obesity  Type 2 diabetes  Cardiovascular disease  Cancer  Kidney disease  Dental disease  Liver disease  Other:______ |
| Mentions children’s health in justification for restrictions | Yes  No |
| Children’s health outcomes cited [select all that apply] | Low diet quality/poor dietary intake  Obesity  Type 2 diabetes  Cardiovascular disease  Cancer  Kidney disease  Dental disease  Liver disease  None stated/not applicable  Other:______ |
| Types of literature cited [select all that apply] | Government report  Peer-reviewed publication  Other report  Website  None  Other: _______ |
| Links to literature cited |  |
| Required waiver contents | Justification  Implementation plan  Education and outreach plan for participants  Evaluation  Other:_____ |
| Implementation timeline | Text box |
| Other reporting requirements | Text box |
| Any other relevant information not captured above | Text box |
